# N-Palmitoyl glycine activates transient receptor potential channel 5 and increases the risk of Brugada syndrome

**DOI:** 10.1101/2024.10.21.24315897

**Authors:** Hongxuan Xu, Bingxun Li, Ying Chen, Yanyun Lin, An Zhang, Lin Wu

**Author notes:** Corresponding author: Lin Wu; address: NO.8 Xishiku Street, Department of Cardiology, Peking University First Hospital, Beijing, China. These authors contributed equally to this study (Hongxuan Xu, Bingxun Li).

## Abstract

**Background:** Brugada syndrome (BrS) is an arrhythmic disorder associated with an increased risk of sudden cardiac death; however, current treatment options are limited due to their side effects and variable efficacy.

**Methods:** In this study, we first employed Mendelian randomization analysis utilizing proteomic, transcriptomic, and metabolomic data to identify potential therapeutic targets for BrS. Ex vivo perfused heart models were used to assess the effects of the potential targets on action potentials and QT intervals. Calcium indicators were employed to evaluate calcium homeostasis in primary cardiomyocytes, and patch-clamp techniques were used to investigate the impact on Nav1.5 and TRPC5 channels.

**Results:** Our findings indicate that N-palmitoyl glycine (PalGly) is linked to an increased risk of BrS and interacts with BrS-associated proteins, demonstrating moderate binding affinities for DCC, CR1, CTSB, NAAA, DEFB1, EPHA1, IGF1/IGFBP3/ALS, and LTA. Electrophysiological experiments showed that although PalGly does not interact with Nav1.5, it enhances calcium sparks in ventricular cardiomyocytes. We determined that the calcium-modulating effect of PalGly is mediated by its binding to and activation of the transient receptor potential channel 5 (TRPC5) channel. Furthermore, PalGly was found to shorten the QT interval and action potential duration in Langendorff-perfused rabbit hearts and isolated rabbit cardiomyocytes. Transcriptomic and lipidomic analyses of PalGly-treated neonatal rat cardiomyocytes revealed significant negative modulation of immune pathways, akin to the effects observed with agonizing TRPC5.

**Conclusion:** Our study underscores the involvement of PalGly, TRPC5, and inflammation-related proteins in the pathophysiology of BrS.

## Introduction

Brugada syndrome (BrS) is a rare arrhythmic disorder with an increased risk of sudden death, characterized by hallmark ST-segment elevation in the right precordial leads of the electrocardiogram (ECG)^1^. BrS is traditionally considered an inherited channelopathy with a Mendelian autosomal dominant pattern. However, the penetrance and causative effect of reported BrS-related genetic variants is often uncertain. The phenotypes of one variant can be discordance between probands and their family members^2^. Current treatments for BrS including antiarrhythmic medication, implantable cardioverter-defibrillator (ICD), and catheter ablation are limited by their side effects and effectiveness^3^. Especially for asymptomatic patients who present typical BrS pattern ECG, these strategies are far from optimal to prevent a likely fatal incidence. In this study, we first use Mendelian randomization (MR) analysis based on proteomics, transcriptomics, and metabolomics to explore potential causal targets for BrS, and then we test our findings with in vitro and ex vivo electrophysiological experiments.

## Methods

### BrS cohort and ECG-wide association study

Summary-level statistics for BrS were obtained from a meta-analysis study comprising 2,820 unrelated cases and 10,001 controls of European ancestry^4^. The diagnosis followed the guidelines from the 2013 Heart Rhythm Society, European Heart Rhythm Association and Asia Pacific Heart Rhythm Society expert consensus statement^5^, the 2015 European Society of Cardiology guidelines^6^ and the 2017 American Heart Association guidelines^7^. Inclusion criteria required a type 1 BrS ECG, characterized by coved-type ST elevation at baseline (spontaneous) or after a drug challenge test, in one or more leads of the right precordial leads (V1 and/or V2), either in the standard (fourth intercostal space) or higher positions (second or third intercostal spaces). Diagnostic ECGs were centrally reviewed by a cardiac electrophysiologist experienced in BrS to confirm that the diagnostic criteria were met. The genetic associations were adjusted for the first six genetic principal components, SNPs that were missing in four or more of the ten strata, as well as those with a heterogeneity test P < 1e-6 were excluded. Top variants of significant exposures were looked up in ECGenetics (http://www.ecgenetics.org)^8^ to explore their effect on R-R adjusted three-lead exercise electrocardiogram (ECG) morphology. The GWAS for ECG morphology consists of comprehensive deep phenotyping of 77,190 ECGs in the UK Biobank across the complete cycle of cardiac conduction, resulting in 500 spatial-temporal data points. Summary-level statistics for QTc consists of 84 630 UK Biobank pariticipants^9^.

### Multi-omics GWAS data source

Summary-level statistics of genome-wide association studies (GWAS) for two protein quantitative trait loci (pQTL) studies were obtained from deCode genetics^10^ and UKB-PPP^11^. Proteomics from deCode genetics consists of 4,907 circulating proteins in 35,559 Icelanders measured by SOMAscan version 4. Proteomics from UKB-PPP consists of 2,923 circulating proteins measured by Olink technology in 54,219 UK Biobank participants with the most European ancestry (around 95%). The protein level of these two studies was inverse rank normalized to perform covariates-adjusted genetic association. In addition, eQTL data was obtained from the eQTLgen Consortium which measured transcriptomics by arrays and RNA-seq from 311,684 blood and PBMC (peripheral blood mononuclear cell) samples (Predominantly European ancestry)^12^. Summary-level statistics of genetic associations with inverse rank normalized levels of 1,091 blood metabolites and 309 metabolite ratios quantified by the Metabolon HD4 platform in 8,299 unrelated European ancestry individuals from the Canadian Longitudinal Study on Aging (CLSA) cohort^13^ were obtained.

### Cardiometabolic traits

A total of 28 cardiometabolic traits were included, including lipids (total cholesterol, triglycerides, high-density lipoprotein cholesterol (HDL-C), low-density lipoprotein cholesterol (LDL-C), apolipoprotein A1 and B (ApoA1, ApoB), and lipoprotein A Lp(a))^14,15^, blood pressure (systolic blood pressure (SBP), and diastolic blood pressure (DBP))^16^, glycemic traits (fasting glucose, fasting insulin, 2-hour post-challenge glucose, and HbA1c)^17,18^, and anthropometric traits (body mass index (BMI), waist circumference, hip circumference, and waist-to-hip ratio)^19^. With 11 cardiometabolic diseases, including atrial fibrillation (AF), stroke and stroke subtypes (large artery atherosclerosis, small-vessel, and cardioembolic)^20^, coronary heart disease (CHD)^21^, heart failure (HF)^22^, aortic aneurysms^23^, chronic kidney disease^24^, type 1 diabetes^25^ and type 2 diabetes^26^. Most participants in these cohorts were of European ancestry (Table S1).

### Exposure instrument variables selection

Genome-wide suggestive SNPs (p < 1e-6) in weak linkage disequilibrium (r^2^ < 0.1, based on the 1000 Genomes Phase 3 European reference panel) within ± 1 Mb from target gene loci associated with the proteins were obtained. Independent effect SNPs for metabolites QTL were identified by genome-wide suggestive threshold (p < 1e-6) and were clumped by a 1Mb distance with a linkage disequilibrium threshold r^2^ = 0.1. Clumping was performed by Plink version 1.9^27^. F-statistics of less than ten were used to exclude SNPs that were weak instruments for a limited proportion of the variance explained.

### MR analysis

The included cohorts consist of almost European ancestry, no individuals likely to overlap. Two-sample MR analysis based on indexed SNPs for circulating proteins, RNAs, and metabolites was used to capture the associations with the risk of BrS. An inverse variance weighted (IVW) random-effects model was employed to estimate the causal effects of genetically proxied targets on BrS. Bonferroni correction was used to account for multiple tests. Weighted median, MR Egger and Steiger test were used to employ sensitivity analysis, heterogeneity test, and causal direction test. All MR analyses were performed in R version 4.3.2 by TwoSampleMR package^28^. Findings were reported according to the STROBE-MR (Strengthening the Reporting of Mendelian Randomization Studies) guidelines^29^.

### Colocalization analysis

Colocalization analysis was employed by the coloc R package^30^ to test whether identified exposures and BrS were associated and shared the same causal variant within ± 1 Mb from target gene loci associated with the proteins. We set prior probabilities of the SNP being only associated with exposure (p1) or outcome (p2) at 1e-4; and the probability of the SNP being associated with both traits (p12) at 1e-5, posterior probability for shared causal variants (PPH4) ≥0.8 were considered to have strong evidence of colocalization. The set of significant genes and proteins with high colocalization support was set forth to an overrepresentation analysis using the pathways described in the Gene Ontology and the KEGG database. Selected pathways were those significantly enriched at an FDR <0.05.

### Mediation analysis

We explored the mediation effect of circulating proteins on BrS via metabolites using the delta method to approximate confidence intervals by the RMediation package^31^. The MR estimate for the mediated effect of genetically predicted levels of circulating metabolites on BrS was divided by the total effect of genetically predicted levels of circulating proteins on BrS risk estimated in the IVW univariable MR, with standard errors estimated using the propagation of error method to calculate mediation proportion.

### Molecular Docking

The 3D structures of DCC, CR1, CTSB, NAAA, DEFB1, EPHA1, IGF1/IGFBP3/ALS, and LTA were obtained from RCSB Protein Data Bank (PDB ID: 2ED7, 2ED8, 2ED9, 2EDB, 2EDD, 2EDE, 2MCY, 2MCZ, 3AI8, 1HUC, 6DXW, 6DXX, 1IJV, 2K1K, 2K1L, 7WRQ, 4MXV, 4MXW) and was subjected to processing through the pdb2pqr before initiating the docking analysis. The entire docking process was conducted using Autodock vina 1.2.0 by Dockey graphical interface^32^ with default parameters. The visualization was performed by PyMOL 3.0.3.

### Isolation of rabbit cardiomyocytes

Rabbit ventricular cardiomyocytes were isolated enzymatically, as previously described^33^. The animal used in this study conformed to the “Guide for the Care and Use of Laboratory Animals” published by the US National Institutes of Health (NIH Publication No. 85-23, revised 2011). The ethical clearance was granted by the Institutional Animal Care and Use Committee of Peking University First Hospital (approved number: J2024090), affirming the conscientiousness of their treatment and welfare. Male New Zealand rabbits weighing 2.5 to 3.0 kg (approximately 3-month-old, Fangyuanyuan, China) were anaesthetized with xylazine (16 mg/kg, Huamu; China) and ketamine (40 mg/kg CAHG; China) (IV) and injected with 5,000 units/kg of heparin. The absence of pain response and corneal reflex served as the indicators to ascertain the appropriate level of anaesthesia depth. Rabbit hearts were rapidly excised and retrogradely perfused through the aorta on a standard Langendorff apparatus for 10 min at a flow rate of 10 mL/min with a calcium-free Tyrode solution continuously gassed with 100% O_2_ at 37 °C. The hearts were perfused in a recirculating manner for 40–60 min (ventricular cardiomyocytes) with 100 mL of calcium-free Tyrode solution containing 60 mg of collagenase (type II, Sigma) and 100 mg of bovine serum albumin (BSA, Sigma). The enzymes were washed out for 5 min via perfusion with a calcium-free Tyrode solution. The ventricles were cut into small chunks, and the tissue was manually pipetted with a disposable plastic transfer pipette for 1-2 min to mechanically separate the cells. The cell and enzyme mixture were filtered through a 200-micron nylon mesh into centrifuge tubes and the cells were stored in KB solution. Cells of approximately the same length and with clear stripes, a rod shape, a clean and smooth surface, and no spontaneous contractions over 1 minute of observation were selected for use in the experiments. The animal use was approved by the Institutional Animal Care and Use Committee of Peking University First Hospital. The isolated cardiomyocytes were then incubated with N-palmitoyl glycine (MedChemExpress) for at least 15 min to study its effect on action potential and calcium spark.

### Isolated Rabbit Heart Assay

Following the excision, the heart was carefully removed and perfused with a specially modified Krebs–Heinsleit (K–H) perfusate (pH 7.4, gassed with 95% O_2_ and 5% CO_2_) that was prewarmed to 37 °C at a controlled speed of 20 mL/min after the aorta was cannulated. The K–H solution contained (in mM) 118 NaCl, 2.8 KCl, 1.2 KH_2_PO_4_, 2.5 CaCl_2_, 0.5 MgSO_4_, 2.0 sodium pyruvate, 5.5 glucose, 0.57 Na2EDTA, and 25 NaHCO_3_. To ensure successful electrical pacing, the right atrium was partially removed and the whole atrioventricular block was induced by thermo-ablating the atrioventricular nodal area. A custom-made bipolar Teflon-coated electrode was placed on the right ventricular septum to pace the heart. Electrical stimuli, 3 ms in width and 3-fold threshold amplitude were delivered to the pacing electrode at a frequency of 1 Hz using an EP-4 stimulator (EP MedSystems, ST. JUDE MEDICAL). After ventricular pacing was initiated, a 30-minute period was allowed for the equilibration of heart rhythm to achieve a stable and consistent rhythm before the subsequent measurements or interventions. Pressure-contact Ag– AgCl MAP electrodes were placed on the epicardial and endocardial ventricular walls, respectively. The ventricular MAP and pseudo-ECG were continuously monitored, amplified, filtered, and digitized in real-time using a BIOPAC Systems MP 150 signal processor and displayed on a computer screen using the AcqKnowledge 4.2 for MP150 software (BIOPAC Systems, Inc.). The duration of the monophasic action potential at 30, 50, and 90% repolarization (MAPD30/50/90) was used to monitor left ventricular repolarization. The MAPD30/50/90 was measured at the steady state following the PalGly perfusion.

### Whole-Cell Voltage-Clamp Recordings of Na_V_1.5 Channels in HEK293T Cells

The HEK293T cells were grown in culture dishes for 24 h, and plasmids expressing Na_V_1.5 (NM_198056) were transfected into HEK293T (National Collection of Authenticated Cell Cultures, China) cells using lipofectamine 3000 (Thermo Fisher Scientific). The extracellular Solution (mM) contained: 137 NaCl, 1 MgCl_2_·6H_2_O, 4 KCl, 1.8 CaCl_2_·2H_2_O, 10 D-Glucose, 10 mM HEPES, adjusted to pH 7.4 with NaOH. The intracellular Solution (mM) contained: 50 CsCl, 60 CsF, 10 HEPES, 20 EGTA, and 10 NaCl, adjusted to pH 7.2 with CsOH. Patchmaster software, coupled with an EPC-10 amplifier (EPC-10, Heka Electronic, Germany), was used to collect and store electrophysiological data. Electrodes (1-3 MΩ) were pulled from 1.5-mm borosilicate glass filaments using a P1000 micropipette puller. The holding potential was set at -120 mV, with a series of square-wave pulses from -120 to -10 mV in 10 mV increments for 8000 ms, followed by a step to -10 mV for 30 ms, and then returning to -120 mV. The data were fitted to the Boltzmann equation to derive the steady-state inactivation curve. The holding potential was set at -120 mV, followed by depolarization to 0 mV for 20 ms to stimulate the resting-state sodium channel current. The voltage was then stepped to the condition pulse voltage corresponding to the half-inactivation voltage (V₁/_2_) for 8000 ms, followed by repolarization to -120 mV for 30 ms. Subsequently, depolarization to 0 mV for 20 ms stimulated the half-inactivated state sodium channel current. Finally, the potential was returned to -120 mV, with current recordings taken every 20 seconds. Once steady-state current values were achieved, cumulative drug application was performed, starting with negative control (0.1% DMSO) followed by 1 μM PalGly.

### Whole-Cell Voltage-Clamp Recordings of TRPC5 Channels in HEK293 Cells

The HEK293T cells (China Infrastructure of Cell Line Resource) were grown in culture dishes for 24 h, and plasmids expressing hTRPC5 (NM_012471) were transfected into HEK293T cells using lipofectamine 3000. The extracellular Solution (mM) contained: 145 NaCl, 4 KCl, 1 MgCl_2_·6H_2_O, 2 CaCl_2_·2H_2_O, 10 D-Glucose, 10 HEPES, adjusted to pH 7.4 with NaOH. The intracellular Solution (mM) contained: 120 Cs-aspartic acid, 20 CsCl, 2 MgCl_2_·6H_2_O, 8.8 CaCl_2_·2H_2_O, 10 EGTA, 10 HEPES, 2 Na_2_-ATP, adjusted to pH 7.2 with CsOH. Once a whole-cell seal is established, the cell membrane potential is clamped at -80 mV. The voltage is stepped from 0 mV to -100 mV and maintained for 1 ms, followed by a ramp to 100 mV for 300 ms, and then repolarized to 0 mV for 50 ms. Data collection is repeated every 10 seconds to observe the effects of the PalGly on the TRPC5 current.

### Whole-cell current recording for ventricular cardiomyocytes

The extracellular Solution (mM) contained: 137 NaCl, 4 KCl, 10 HEPES, 10 D-Glucose, 1 MgCl_2_·6H_2_O, 2 CaCl_2_·2H_2_O, adjusted to pH 7.4 with NaOH. The intracellular Solution (mM) contained: 135 KCl, 2 MgCl_2_·6H_2_O, 10 HEPES, 1 EGTA, 4 Mg-ATP, 0.3 NaGTP, adjusted to pH 7.2 with KOH. All the experiments were performed at a temperature of 23 ± 1 °C. For ventricular cardiomyocytes, the cells are clamped at -80 mV, and the recording is switched to the current clamp mode with I = 0. Ventricular myocytes require stimulation to generate action potentials. The parameters for the inward current pulse stimulation are as follows: duration of 10 ms and stimulation amplitude ranging from 0.1 to 1 nA.

### Measurements of Ca^2+^ sparks

Isolated cardiomyocytes were subjected to a fluorescent calcium indicator, Fluo-4-AM (Invitrogen), for 15 minutes at room temperature. Following this incubation period, the cells underwent centrifugation, after which the supernatant was carefully removed. The cells were then resuspended in fresh Tyrode solution containing 250 μmol/L probenecid sodium for an additional 20 minutes to facilitate the de-esterification of the dye and was carried out in laminin-coated dishes. Cytosolic Ca2+ levels were monitored utilizing a confocal microscope (Leica SP8; Leica, Wetzlar, Germany) operating in line-scan mode at a frequency of 700 Hz. Ca^2+^ sparks were invoked after a 20-second field stimulation (1Hz, 4ms, 40V) with a stimulator (MyoPacer EP, IONOPTIX). The subsequent detection and quantification of calcium sparks were performed by SparkMaster 2^34^.

### Lipidomic and transcriptomic analysis

To extract cardiomyocytes, using ice-cold ethanol to anesthetize neonatal rats (0-3 days) followed by cervical dislocation for euthanasia. Dissect immediately the chest cavity to remove the heart. Place the heart in a petri dish containing cold PBS buffer. Cut the heart into small pieces and isolate cardiomyocytes using a neonatal Cardiomyocyte Isolation Kit, rat (Miltenyi Biotec, Germany) according to its protocol. After 1 hour of differential adherence, the suspending cells were considered cardiomyocytes and were transferred to culture flasks containing DMEM supplemented with 20% FBS for continued culture over 48 hours. Following this, the medium is replaced with serum-free DMEM for 24 hours, after which 1 μM PalGly (dissolved in DMSO) or an equivalent volume of DMSO is added for an additional 24-hour incubation. Subsequently, the culture medium is extracted for lipidomic analysis, and RNA is extracted for RNA-seq transcriptomic analysis. The RNA-seq data can be accessed in the GEO database (GSE278421). We analyzed the altered lipid-related pathways by a lipid pathway enrichment analysis (LIPEA). DEGs and GSEA were analyzed using clusterProfiler 4.10.1.

### Statistical analysis

Data from the same cells or hearts before and after treatment are analyzed using a paired samples t-test, while comparisons between different cell groups are conducted using an two-sided independent samples t-test. The concentration-response curves were fitted to a four-parameter Hill equation. All analyses were performed in R 4.3.2 and GraphPad Prism 10.2.3.

### Ethics

The UKB participants gave informed written consent, and ethical approval was obtained from the North West Centre for Research Ethics Committee. The deCode participants who donated samples gave informed consent, and the National Bioethics Committee of Iceland approved the study. The All GWAS human participants provided written informed consent, and all studies had received approval from the appropriate ethical review boards.

## Results

The flowchart of the MR design is presented in **Figure 1**. Instrument variables used in this study are presented in **Table S2-5**.

**Figure 1.**
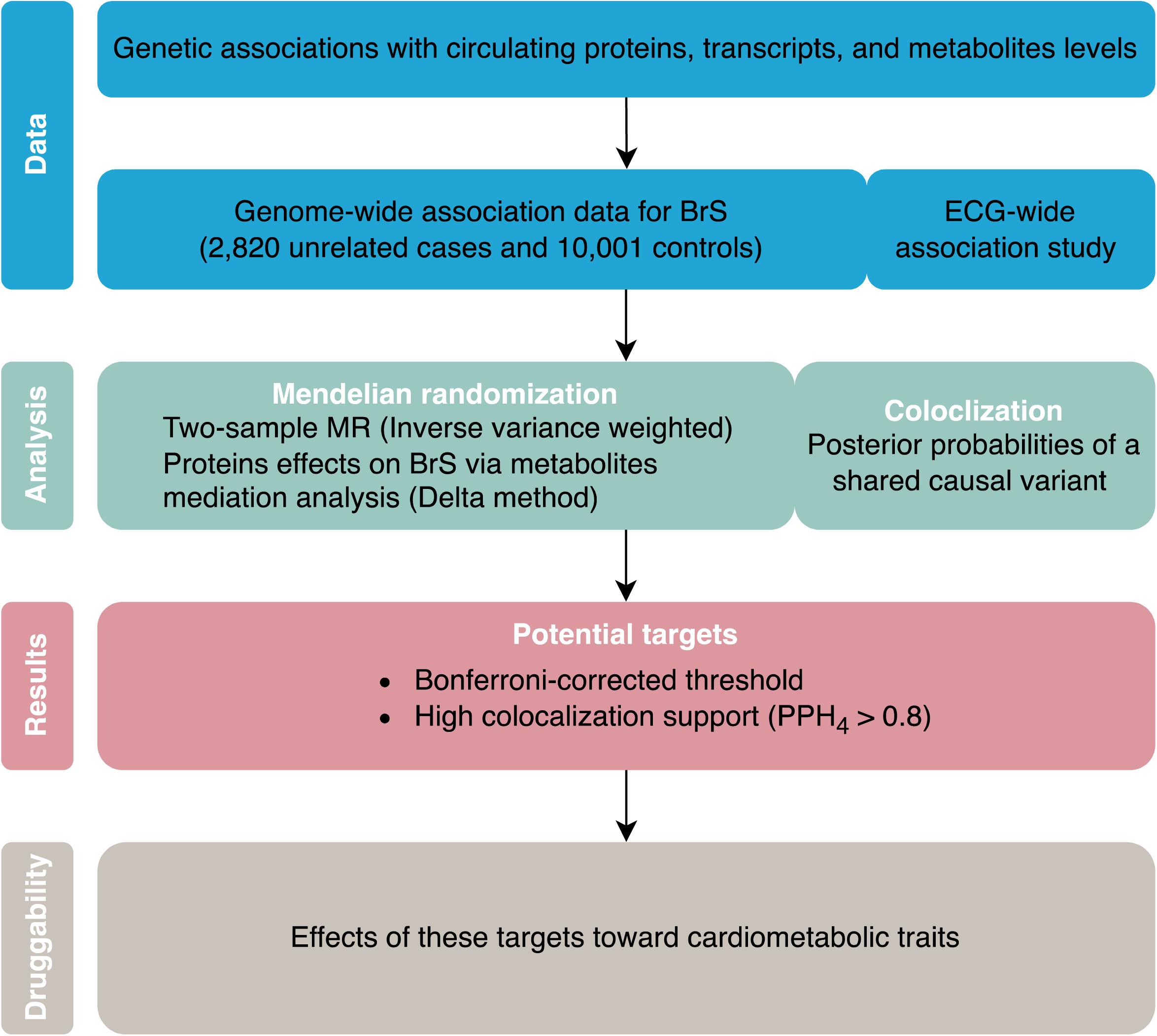
Flowchart of the MR design

### Circulating transcripts and proteins that causally associated with the risk of BrS

The overview of the results of MR and colocalization analysis is presented in **Figure 2**. A total of nine proteins (H6PD, ANXA4, CXCL6, LRP11, CTSB, DEFB1, PDGFD, SPATA20, GSS) from deCode genetics reached the Bonferroni corrected threshold (p < 0.05/1320) among which four proteins (CXCL6, CTSB, SPATA20, DEFB1) were with high colocalization support (PPH_4_ > 0.8) (**Table 1**), while 13 proteins (CR1, CXCL6, NAAA, ARSB, HCG22, LTA, CTSB, IGFBP3, EPHA1, ECHDC3, AAMDC, ZFYVE19, DCC) from UKB-PPP reached Bonferroni corrected threshold (p < 0.05/ 1590) among which eight proteins (CR1, CXCL6, NAAA, LTA, IGFBP3, EPHA1, AAMDC, DCC) were with high colocalization support. In addition, 12 genes at the transcript level with high colocalization support from eQTLgen were significantly associated with BrS (LUZP1, PCYOX1, NEIL2, MFHAS1, FAM86B3P, SCAND2P, C15orf26, ZSCAN2, ADAMTS17, UBTF, DESI1, EP300). Detailed results are presented in **Table S6-9**. Enrichment analysis showed that IGFBP3 and EPHA1 were significantly associated with fibronectin-binding (GO:0001968) after FDR adjustments.

**Figure 2.**
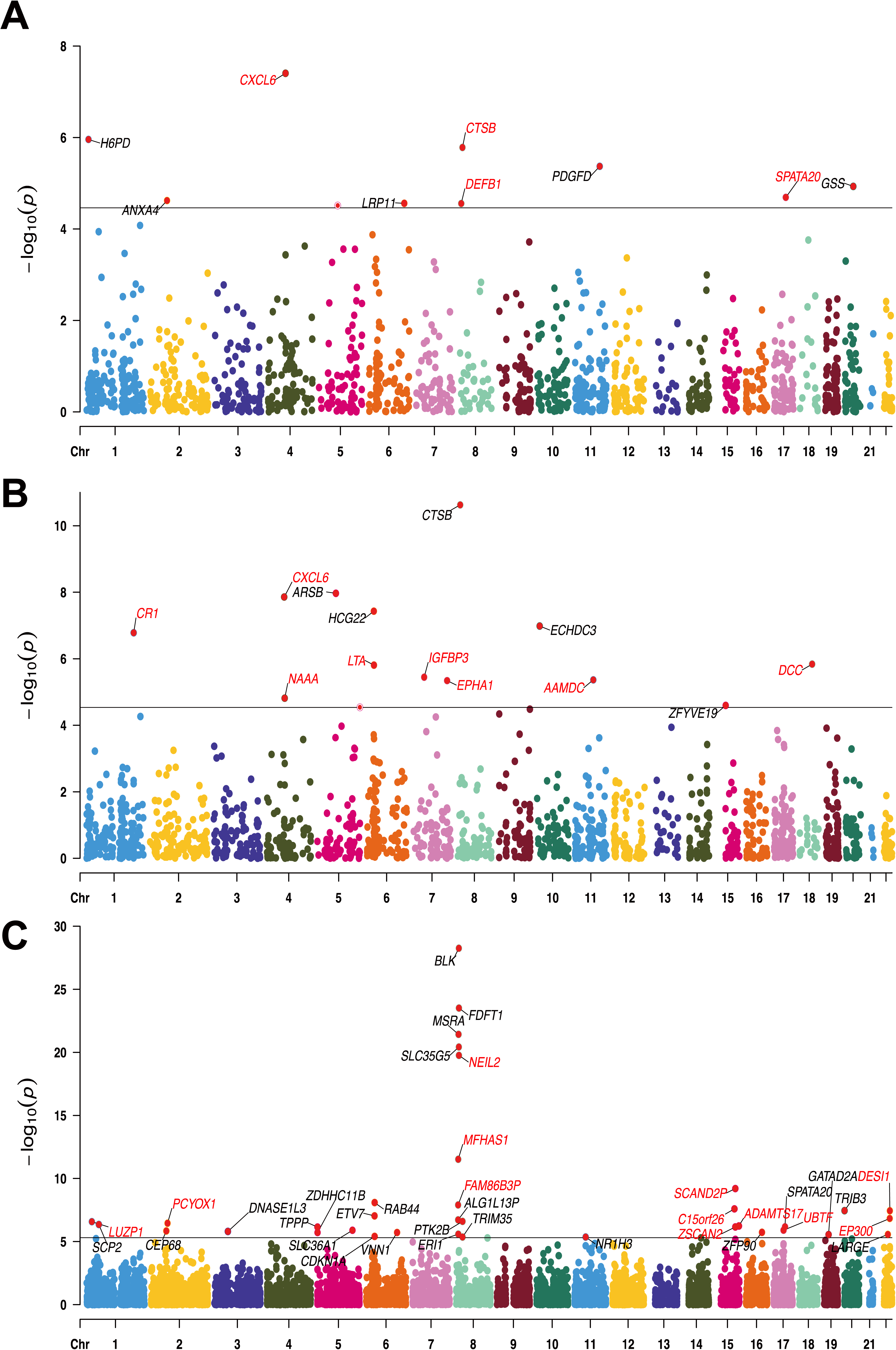
Manhattan plot of significant proteins and transcripts. Red symbols indicate high colocalization support. (A) Proteins from deCode. (B) Proteins from UKB-PPP. (C) Transcripts from eQTLgen.

**Table 1.**
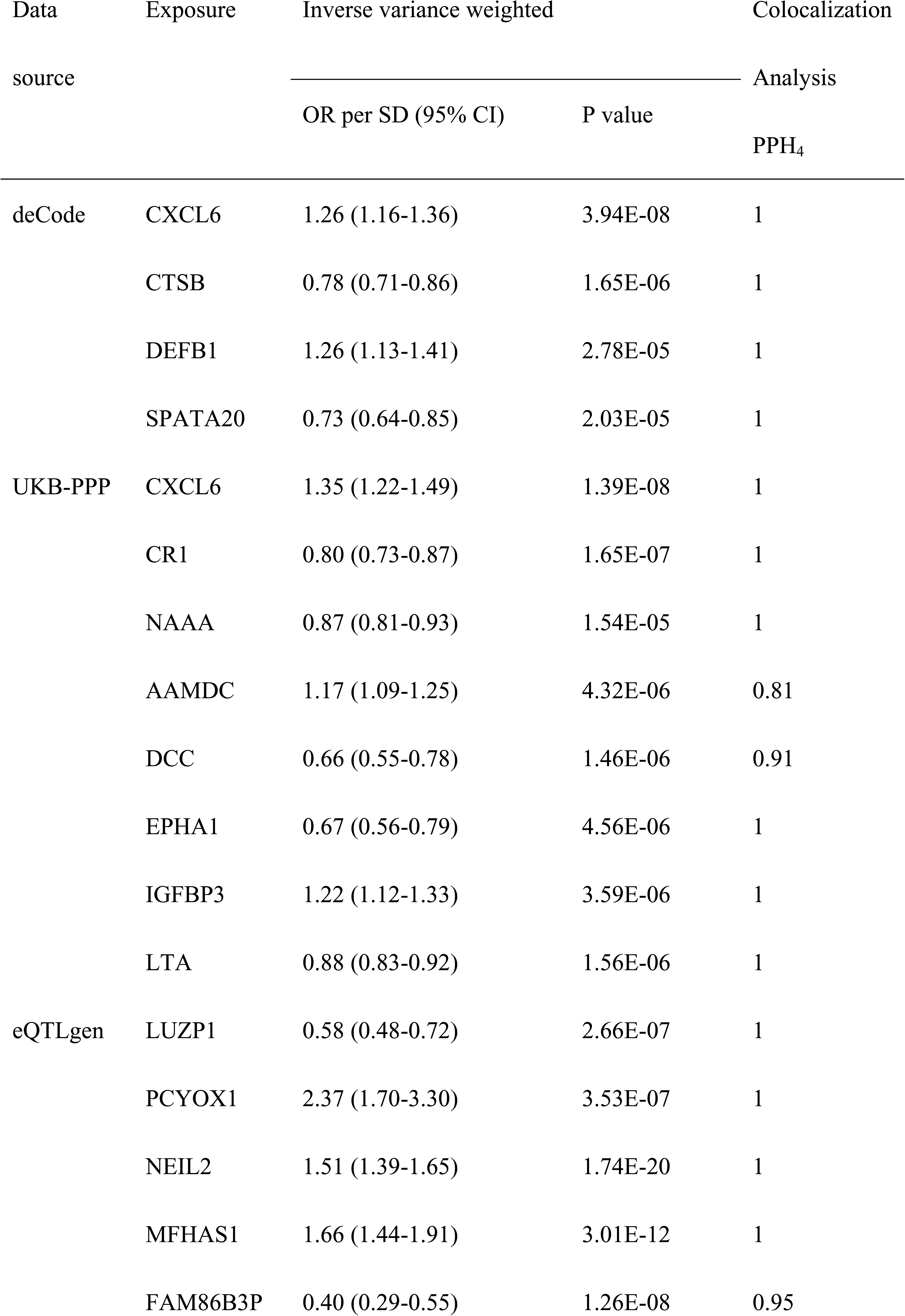
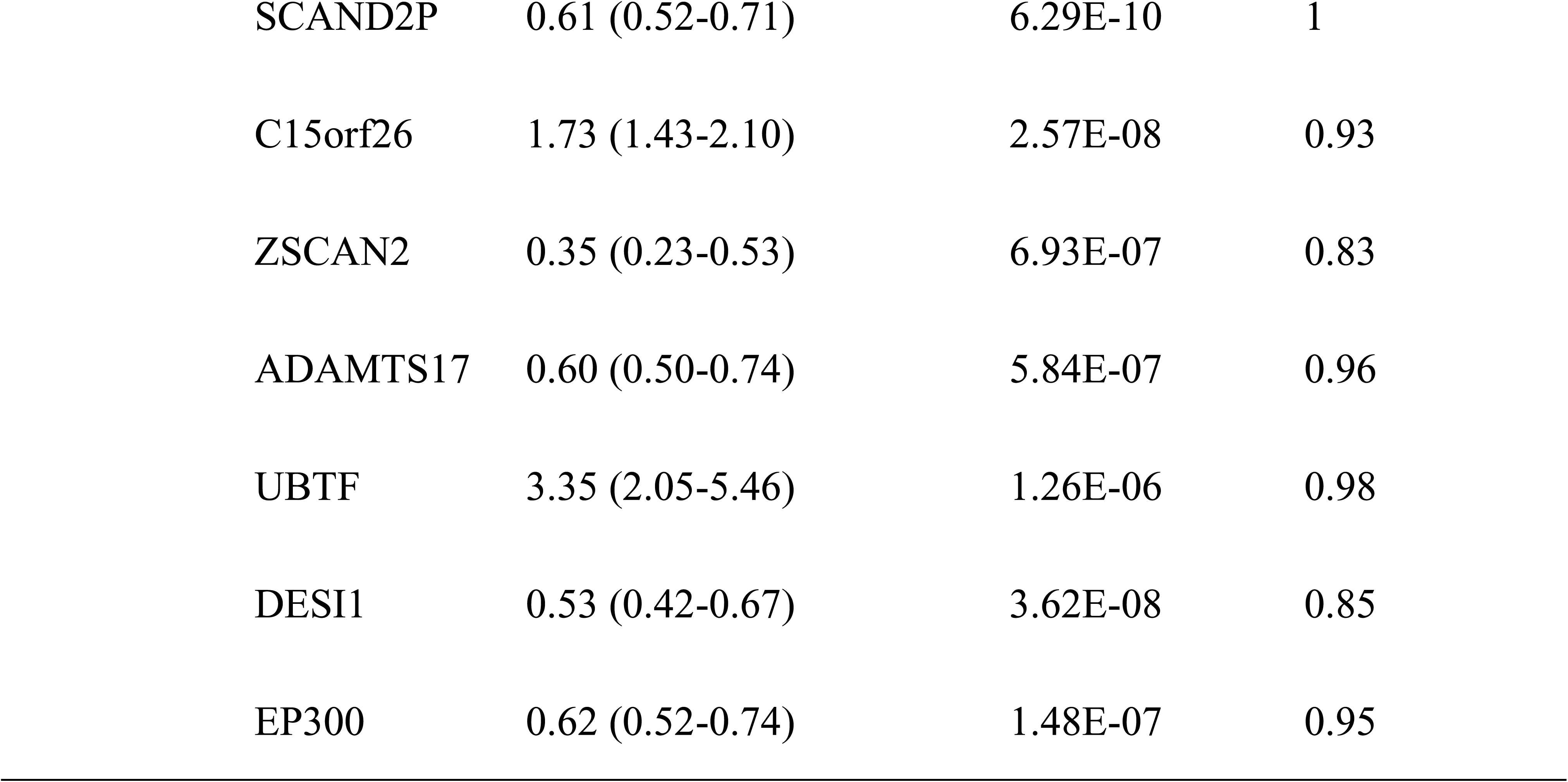
Mendelian randomization analysis and colocalization of circulating proteins and transcripts on Brugada syndorme.

### 3.2 Circulating protein levels affect genetically proxied ECG morphology

Despite the main ECG characteristics of BrS present in the right precordial leads, causal inference of circulating proteins on three-lead exercise ECG morphology GWAS may unveil potential mechanisms. For proteins that passed both MR and colocalization thresholds, top cis-pQTLs were selected to represent the expression of the specific proteins (Figure S1). The AAMDC, CXCL6, CTSB, and NAAA affected the slope of the ST segment, higher expression of CXCL6 increased the risk of BrS while causing ST-segment elevation. The SPATA20 was associated with a shift of P wave and PR interval, we postulate that this may be related to the cis-pQTL rs34081637 located at the CACNA1G locus (adjacent downstream of SPATA20). The voltage-dependent calcium channel, T-type, alpha 1G subunit (encoded by CACNA1G) is one of the components of the calcium clock. Unfortunately, we were unable to identify any pQTLs or eQTLs for CACNA1G to proxy its effects on ECG morphology or BrS.

### 3.3 IGFBP3 increases the risk of BrS and is mediated by N-palmitoyl glycine

A total of 69 circulating metabolites were nominally significant, and only one metabolite (N-palmitoyl glycine, PalGly) reached the Bonferroni corrected threshold (p < 0.05/1190) (**Table S9**). Genetically proxied circulating PalGly significantly increased the risk of BrS (per SD, OR 1.42, 95% CI 1.20-1.67). Mediation analysis showed that one causally BrS-associated circulating protein (IGFBP3) increased the risk of BrS partly via PalGly (Table 2).

**Table 2.** The mediation effect of PalGly affects BrS.

| Protein | Total effect | Direct effect | Direct effect | Mediation |  | Mediation |
| --- | --- | --- | --- | --- | --- | --- |
|  |  | A | B | effect |  | proportion |
| | $\beta$ (95% CI) | $\beta$ (95% CI) | $\beta$ (95% CI) | $\beta$ (95% CI) | p | |
| IGFBP3 | 0.20 (0.12 to 0.28) | 0.09 (0.05 to 0.12) | 0.35 (0.18 to 0.51) | 0.03 (0.01 to 0.04) | 2.25e-06 | 3.05 (0.00 to 24.91) |

### 3.4 PalGly, IGFBP3, CR1, and DCC reduce the risks of cardiometabolic diseases while CXCL6 increases the risks

To determine potential therapeutic targets for BrS, we performed MR analyses of the genetically proxied PalGly and proteins on 28 cardiometabolic traits. Surprisingly, PalGly reduced the risks of AF and HF, which was in discordance with its effect on BrS (Table 3). The IGFBP3 showed similar effects with the PalGly with reduced cardiometabolic ischemic stroke risk (Table 4). The CXCL6 increased the risk of AF while CR1 and DCC reduced the risk of cardioembolic strokes and HF respectively. The effects of the CXCL6, CR1, and DCC toward the cardiometabolic traits were in concordance with their effects towards BrS, indicating these targets were therapeutic. Detailed results of potential targets on cardiometabolic traits were presented in **Table S10**.

**Table 3.**
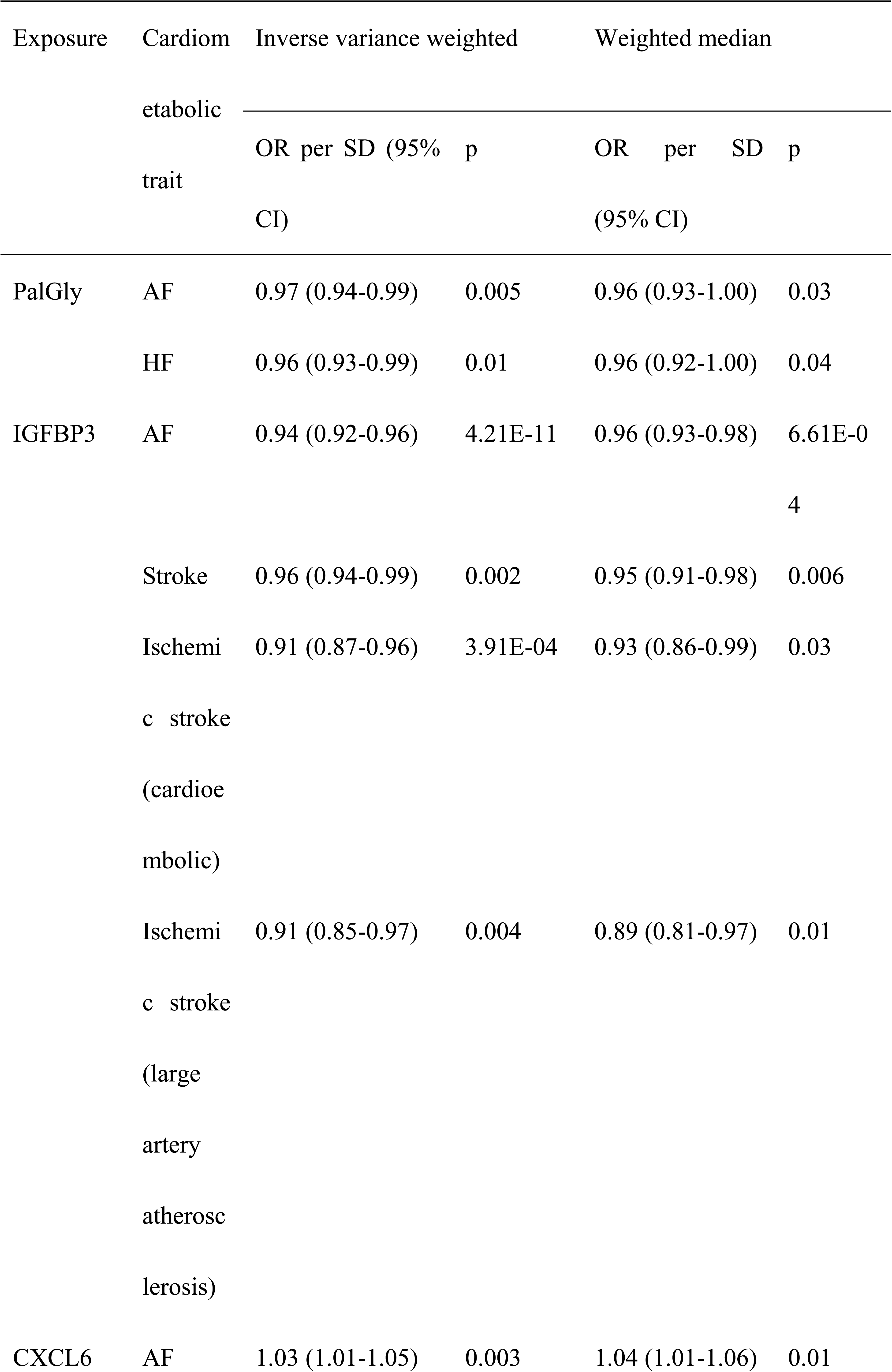
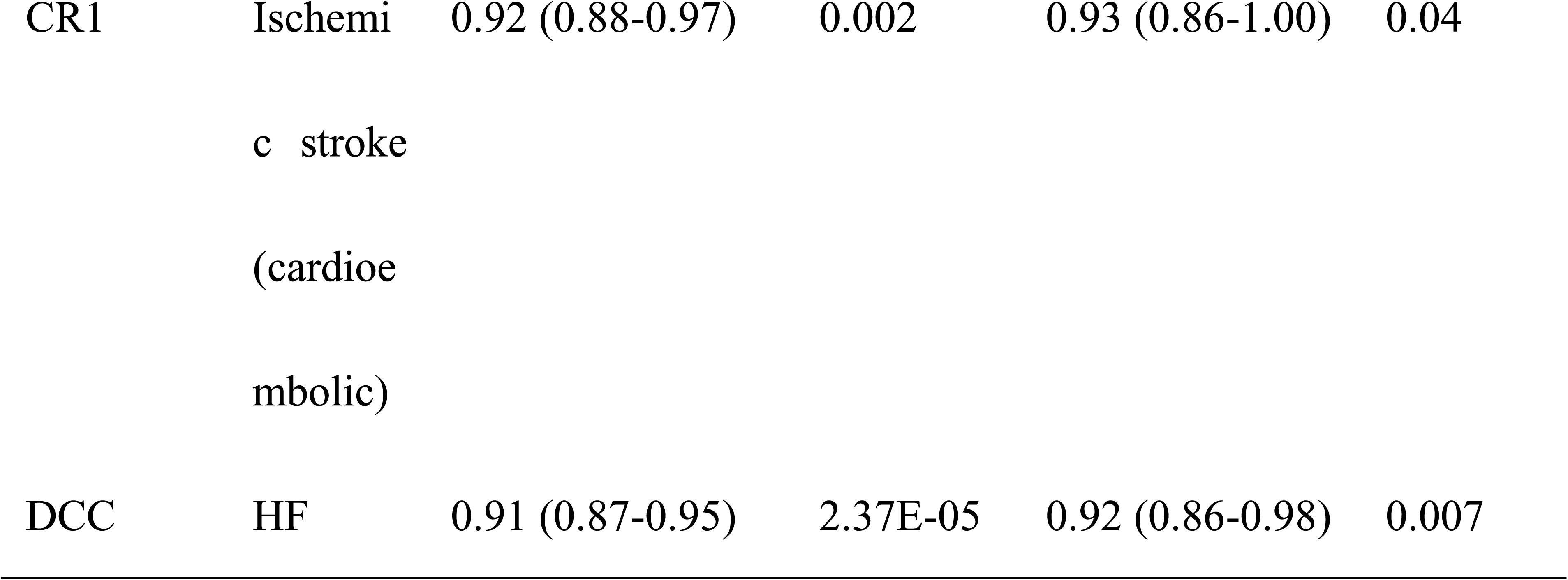
Mendelian randomization analysis of PalGly, IGFBP3 CXCL6, CR1, and DCC on cardiometabolic diseases.

**Table 4.**
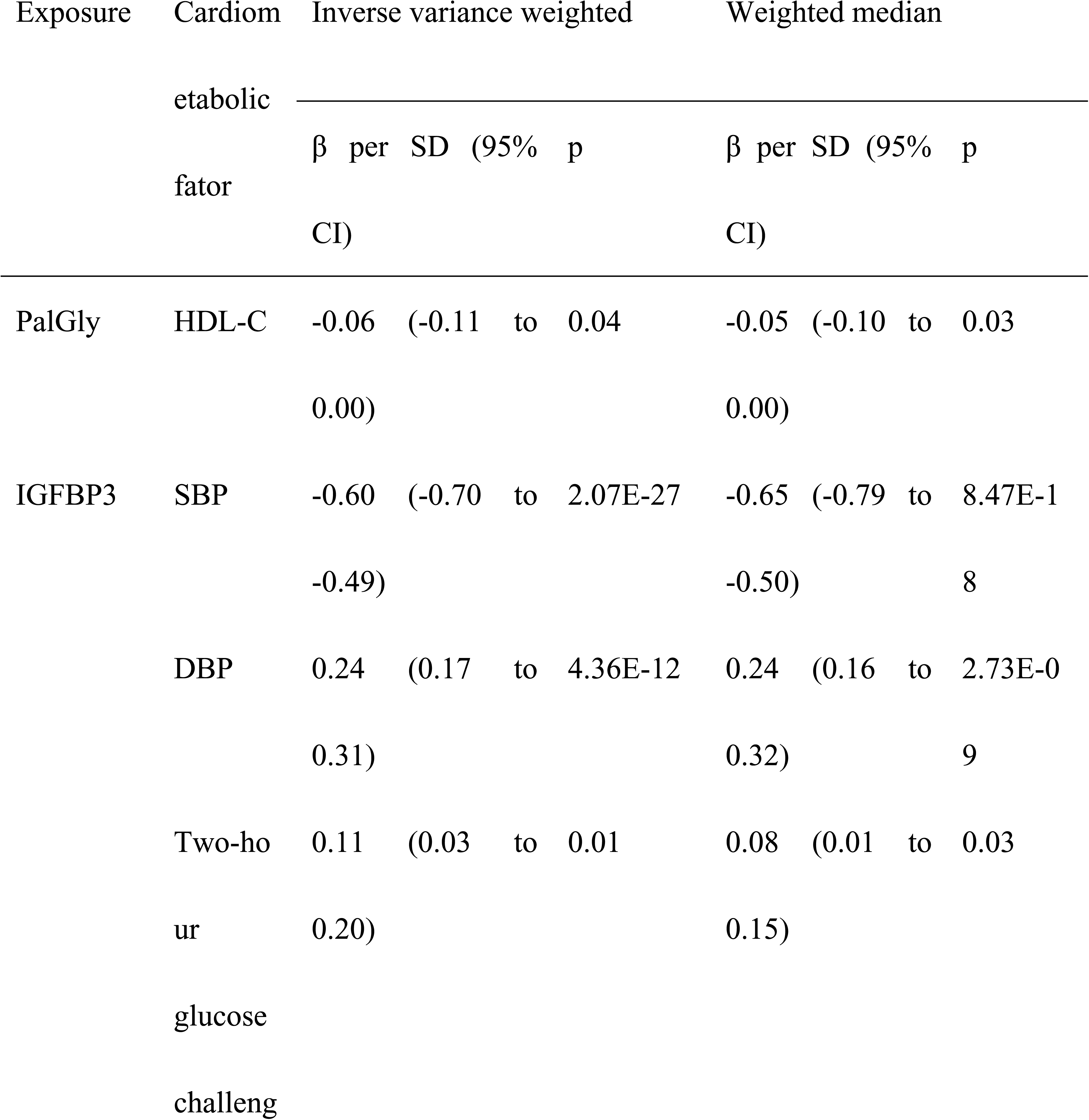
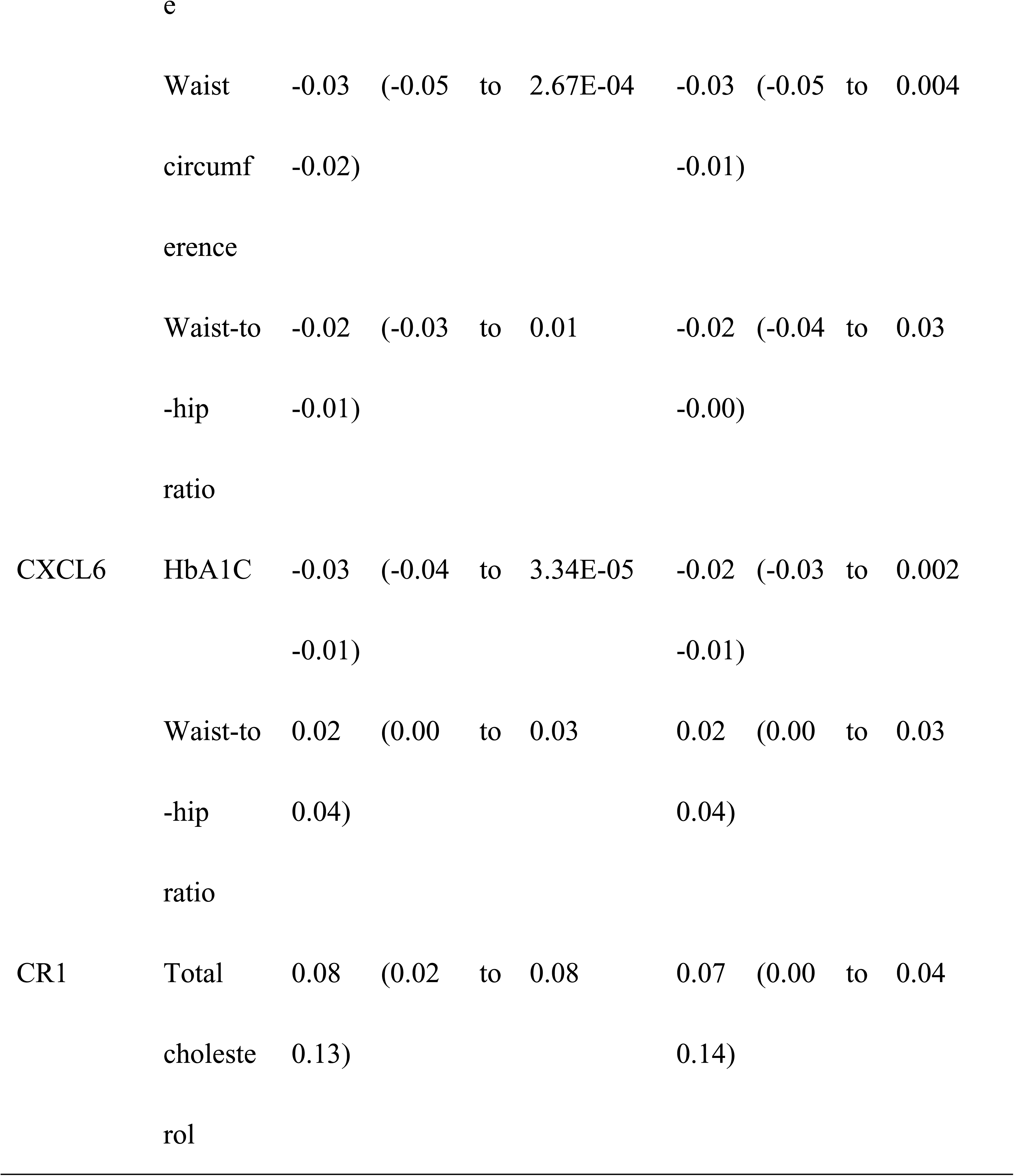
Mendelian randomization analysis of PalGly, IGFBP3, CXCL6, and CR1 on cardiometabolic factors.

### 3.5 PalGly may directly bind to BrS-associated proteins

We performed molecular docking of the PalGly with target proteins. The max binding affinities obtained were -5.4 kcal/mol for DCC, -5.8 kcal/mol for CR1, -5.9 kcal/mol for CTSB, -7.6 kcal/mol for NAAA, -5.8 kcal/mol for EPHA1, -5.7 kcal/mol for IGF1/IGFBP3/ALS complex, and -6.3 kcal/mol for LTA, indicating that the PalGly has the moderate predicted interaction with these targets (**Table S11**). Docking results also revealed that the PalGly forms three hydrogen bonds and exabits three hydrophobic interactions with DCC, forms three hydrogen bonds and exabits four hydrophobic interactions with CR1, forms two hydrogen bonds and exabits seven hydrophobic interactions with EPHA1, and forms one hydrogen bond and exabits five hydrophobic interactions with IGF1/IGFBP3/ALS complex, suggests stable binding conformations.

### 3.6 PalGly does not interact with Nav1.5 but increases calcium sparks of ventricular cardiomyocytes

Mutations in the SCN5A gene are the most known in BrS, therefore we first investigated whether PalGly administration under maximum solubility (1 μM) can affect peak sodium current (Na_v_1.5), and results showed that the effect was not significant (**Figure 3A**). Next, because the PalGly can induce calcium influx in neurons^35^, we investigate its effect on calcium spark on rabbit primary ventricular cardiomyocytes. After 1 μM PalGly infusion and at least 20-second 1 Hz field stimulations, the amplitude of calcium sparks was significantly increased (**Figure 3B**) with increased width, duration, and frequency, while the time to peak of sparks was significantly reduced.

**Figure 3.**
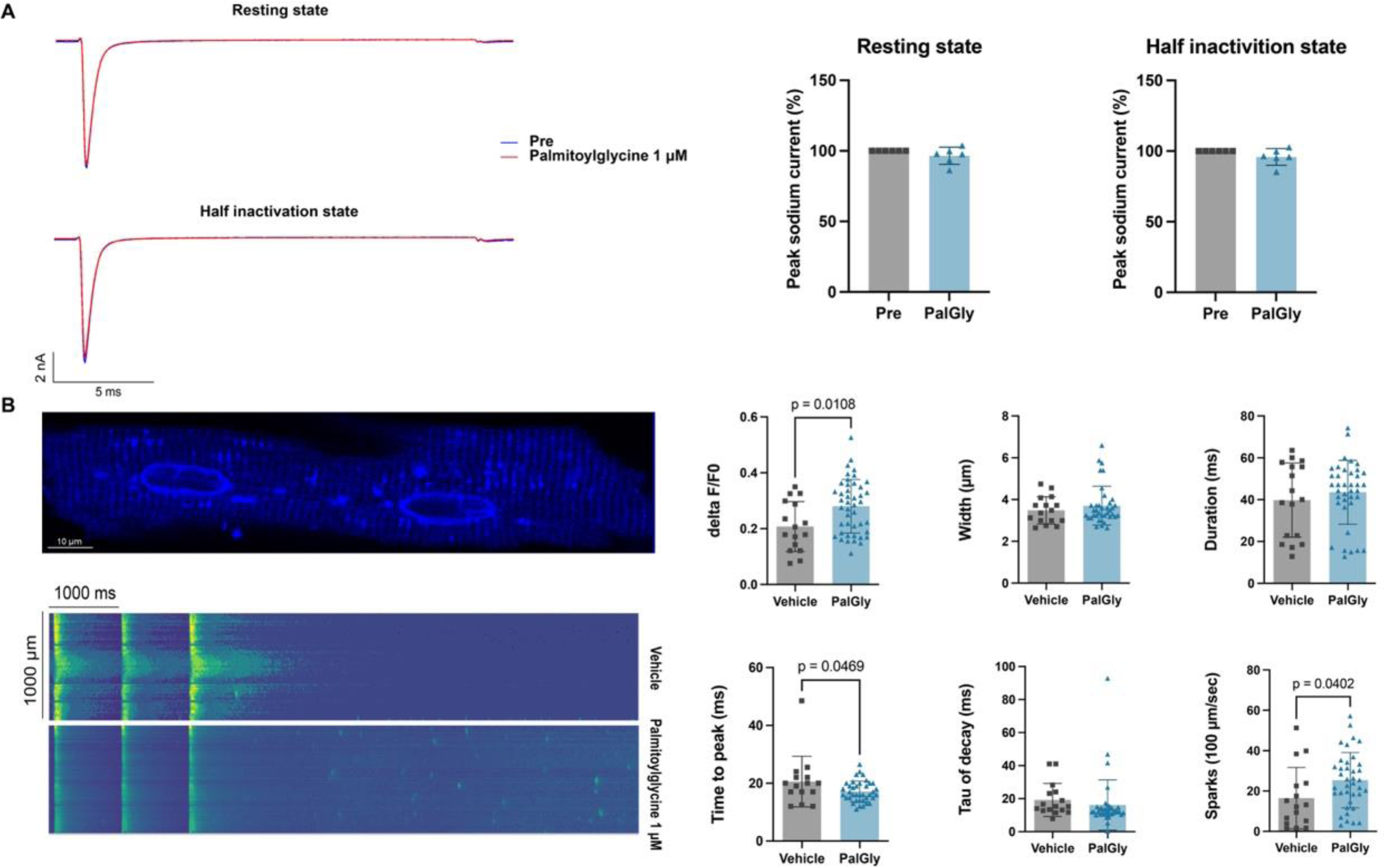
Sodium current and calcium spark traits induced by the palmitoylglycine. (A) Left: representative peak sodium current (Na_v_1.5) before and after the PalGly administration in resting state or half inactivation state. The maximum solubility of palmitoylglycine in extracellular fluid is around 1 µM. Right: normalized current before and after PalGly (n = 5, paired t-test). (B) Left top: representative image of calcium in primary ventricular cardiomyocytes dyed by Fluo4, AM. Left bottom: representative image (standard deviation transformed) of calcium sparks after 20-second 1 Hz filed stimulations. Right: Delta F/F0 representing the amplitude of calcium sparks is significantly higher after PalGly administration (nControl = 11, nPalGLy = 38 from 5 rabbits, unpaired t-test), each dot represents a mean value of calcium sparks of one line-scanned ventricular cardiomyocyte. PalGly: palmitoylglycine.

### 3.7 PalGly activates the hTRPC5 channel

PalGly activates a TRP-like channel in dorsal root ganglia^35^, and there is speculation that PalGly is a direct transient receptor potential (TRP) channel 5 (TRPC5) agonist^36^. Therefore, we performed molecular docking that showed PalGly can bind both class 1 & 2 hTRPC5 channels. The docking results for class 1 hTRPC5 (the dominant one) were presented in **Figure 4A**, the binding affinity for class 2 hTRPC5 was -5.9 kcal/mol, and PalGly forms two hydrogen bonds in Chain A (Gln507, Trp435) and exhibits five hydrophobic interactions (Chain A: Ile484, Leu491, Leu514; Chain D Thr603, Thr607). Next, we expressed hTRPC5 in HEK293 cells to perform whole-cell patch-clamp recording. The results demonstrated that PalGly significantly increased both inward and outward currents of hTRPC5 at its maximum solubility (1μM), with the outward current increasing by approximately two-fold and the inward current increasing by approximately six-fold, and these currents can be diminished by the TRPC5 inhibitor ML204 (**Figure 4B**). Since 1 µM reached the maximum solubility of PalGly, it was not possible to calculate the EC50 value for PalGly activating hTRPC5. Consequently, we calculated the EC50 value for PalGly to be approximately 104 nM (inward current) and 128.3 nM (outward current) by co-administering 20 µM of the TRPC5 selective agonist rosiglitazone (**Figure 4C**).

**Figure 4.**
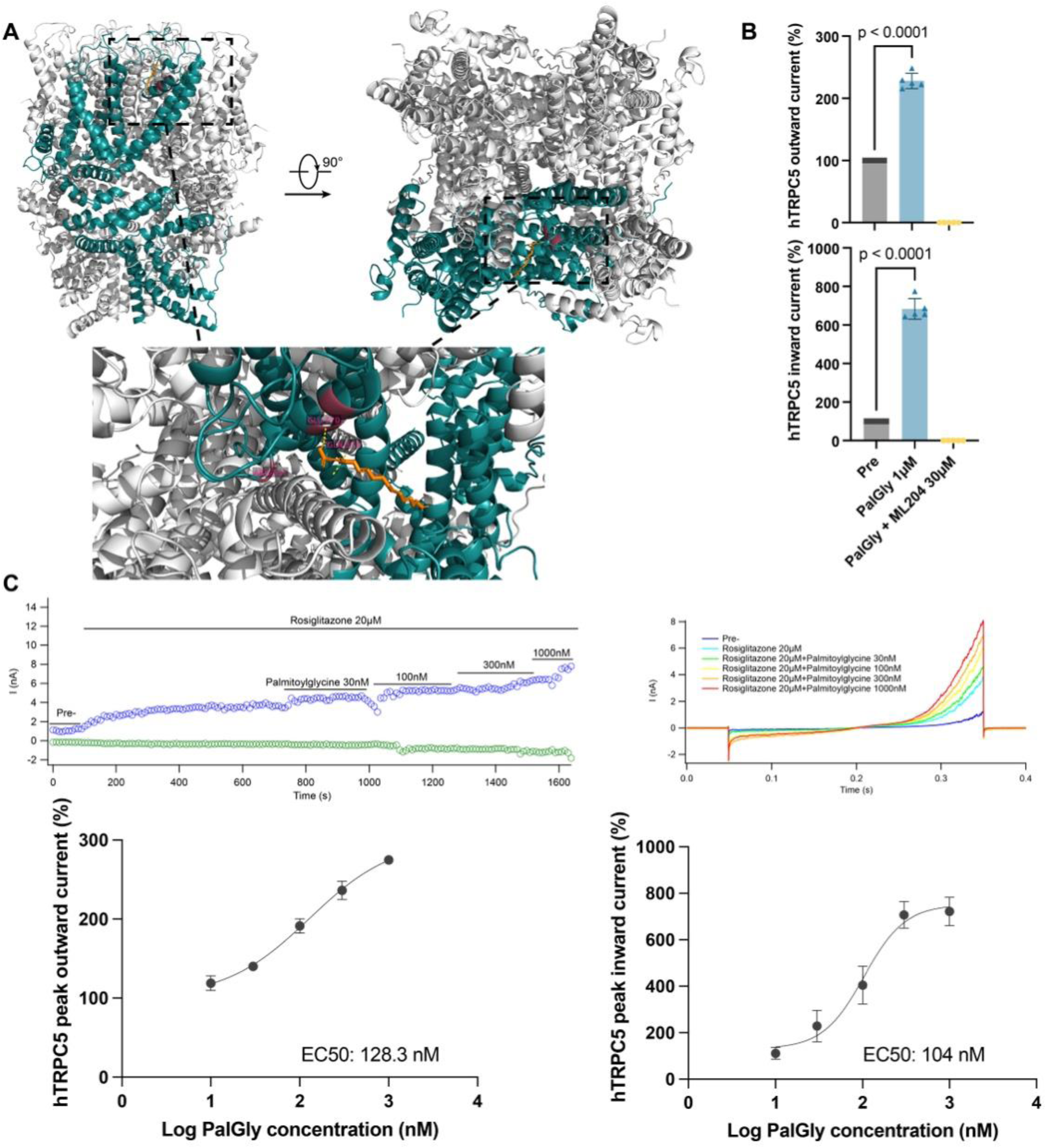
PalGly binds to and agonizes the hTRPC5 channel (A) molecular docking result for PalGly and class 1 hTRPC5. Chain C of hTRPC5 is in cyan, PalGly is in orange, and hydrogen bonds are in magentas. The binding affinity is -5.9 kcal/mol, 3 hydrogen bonds (Gln503, Trp435, Asp433) are formed between PalGly and Chain C of hTRPC5, and 10 hydrophobic interactions (Chain C: Trp435, Met438, Ala441, Ile484, Ile487, Leu491, Leu514, Leu488; Chain D: Thr603, Thr607) are formed. The normalized agonizing effect of PalGly is presented in (B), ML204 is a selective TRPC4/TRPC5 channel inhibitor (≥ 3 cells from 3 dishes, paired t-test). (C) Normalized stimulation curve of PalGly combing rosiglitazone, a TRPC5 activator (≥ 5 cells from 3 dishes). The top two panels are representative recordings of hTRPC5 current, outward current is in blue, and inward current is in green.

### 3.8 PalGly shortens action potential duration and QT interval

We subsequently investigated the effects of PalGly on isolated primary rabbit ventricular myocytes and Langendorff-perfused hearts. After administering or perfusing 1 μM PalGly for at least 10 minutes, we observed a significant shortening in the action potential duration (APD) of both cardiomyocytes (**Figure 5A**) and the overall heart, as well as a significant shortening of the QT interval (**Figure 5B**). Notably, the influence of PalGly appeared to be more pronounced on early repolarization, as indicated by a greater difference in APD30/MAPD30 compared to APD90/MAPD90. Given that rosiglitazone can suppress several cardiac transmembrane ion currents^37^, we did not use it as a TRPC5 positive control. We further assessed the impact of PalGly on QTc through MR analysis, confirming a causal relationship between PalGly and a shortened QTc interval (per SD, β -0.54, 95% CI -1.01, -0.07, p = 0.03).

**Figure 5.**
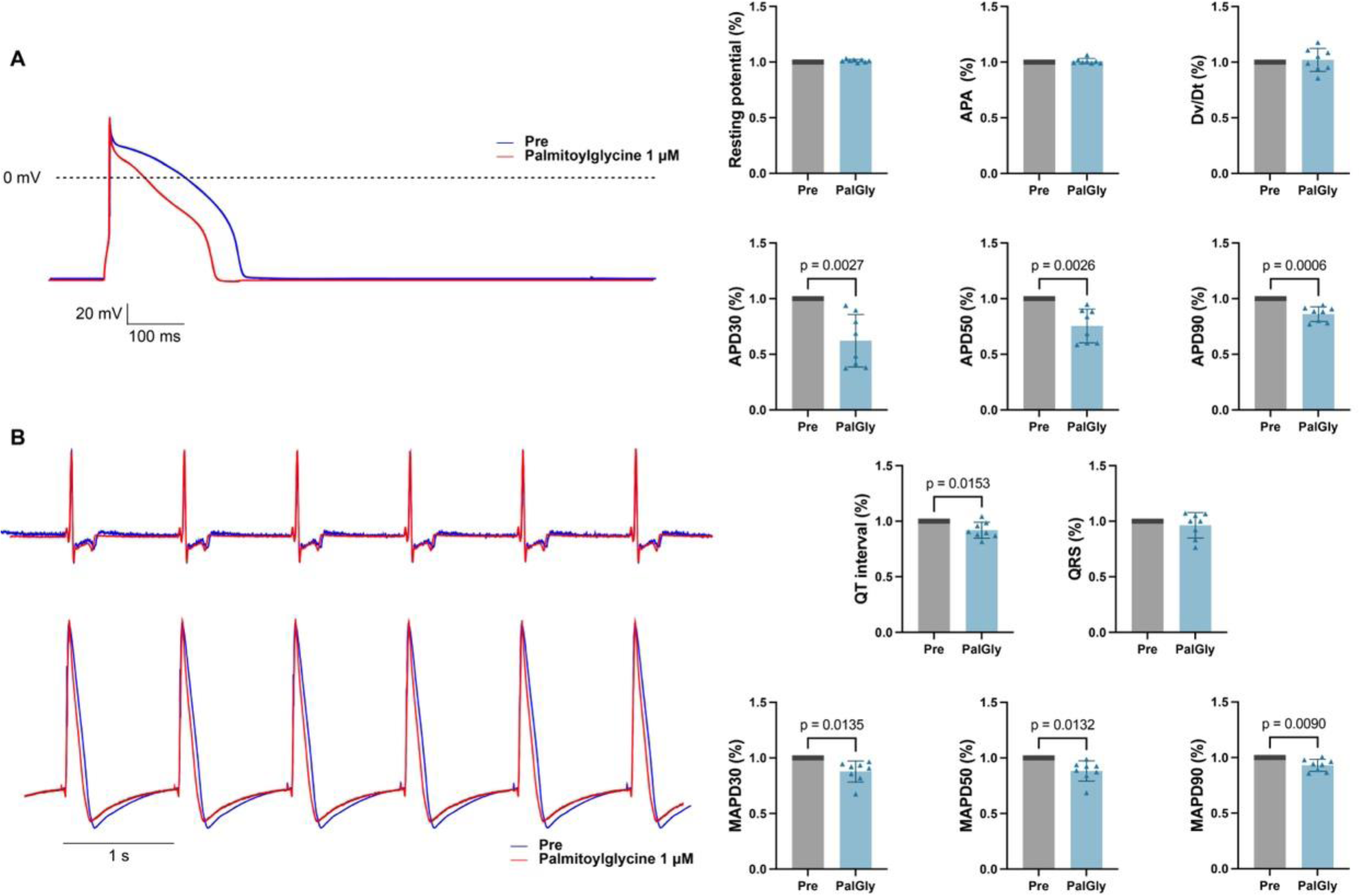
PalGly shortens action potential duration and QT interval (A) APD traits of isolated primary rabbit ventricular cardiomyocytes before and after PalGly administration (n = 8 from 3 rabbits, paired t-test), representative recording is on the left. (B) ECG and APD traits of isolated Langendorff-perfused rabbit hearts (n = 8, paired t-test), representative ECG recording is on the top left and APD recording on the bottom right. APA: action potential amplitude. APD: action potential duration.

### 3.9 PalGly alters the transcriptomic and lipidomic profile of cardiomyocytes

We added a concentration of 3000 ng/ml of PalGly to the culture medium of primary neonatal rat cardiomyocytes for 24 hours. Mass spectrometry quantification of the culture medium revealed that the PalGly concentration decreased to 20.18 ± 16.68 ng/ml, while in the medium of normally cultured cardiomyocytes, the PalGly concentration was approximately 0.24 ± 0.16 ng/ml. Next, following administration with 1 μM PalGly for 24 hours, we extracted the culture medium to perform non-targeted lipidomic analysis, and RNA for RNA-seq and subsequent transcriptomic analysis. Five lipid species were significantly reduced (LPC 18:1, PA 33:1; PA 16:0-17:1, CL 68:3; CL 34:1-34:2, FA 19:1, and PC 32:4e;), while 15 lipid species were significantly increased (FA 20:2, FA 22:3, PG 34:0; PG 16:0-18:0, PE 36:1, FA 18:1, FA 18:2, LPE 16:1e, LPE 18:2e, PI 40:5; PI 18:0-22:5, PI 38:5; PI 18:1-20:4, SM d44:3, PS 40:6, PE 40:5; PE 18:0-22:5, PE 40:4, and TAG 58:5e; TAG 18:0e-18:1-22:4) (**Figure 6A**). Enrichment analysis indicated that these lipid alterations were associated with glycerophospholipid metabolism, ferroptosis, and autophagy (**Figure 6B**). The differential expression genes (DEGs) from RNA-seq are presented in **Figure 6C**. Gene Ontology (GO) and KEGG enrichment revealed that PalGly not only induces changes in autophagy-related genes but is also associated with alterations in various pathways, including ubiquitin-related pathways, MAPK, and TNF signalling (**Figure 6D**). Additionally, Gene Set Enrichment Analysis (GSEA) indicated that PalGly negatively regulates the innate immune response (**Figure 6E**).

**Figure 6.**
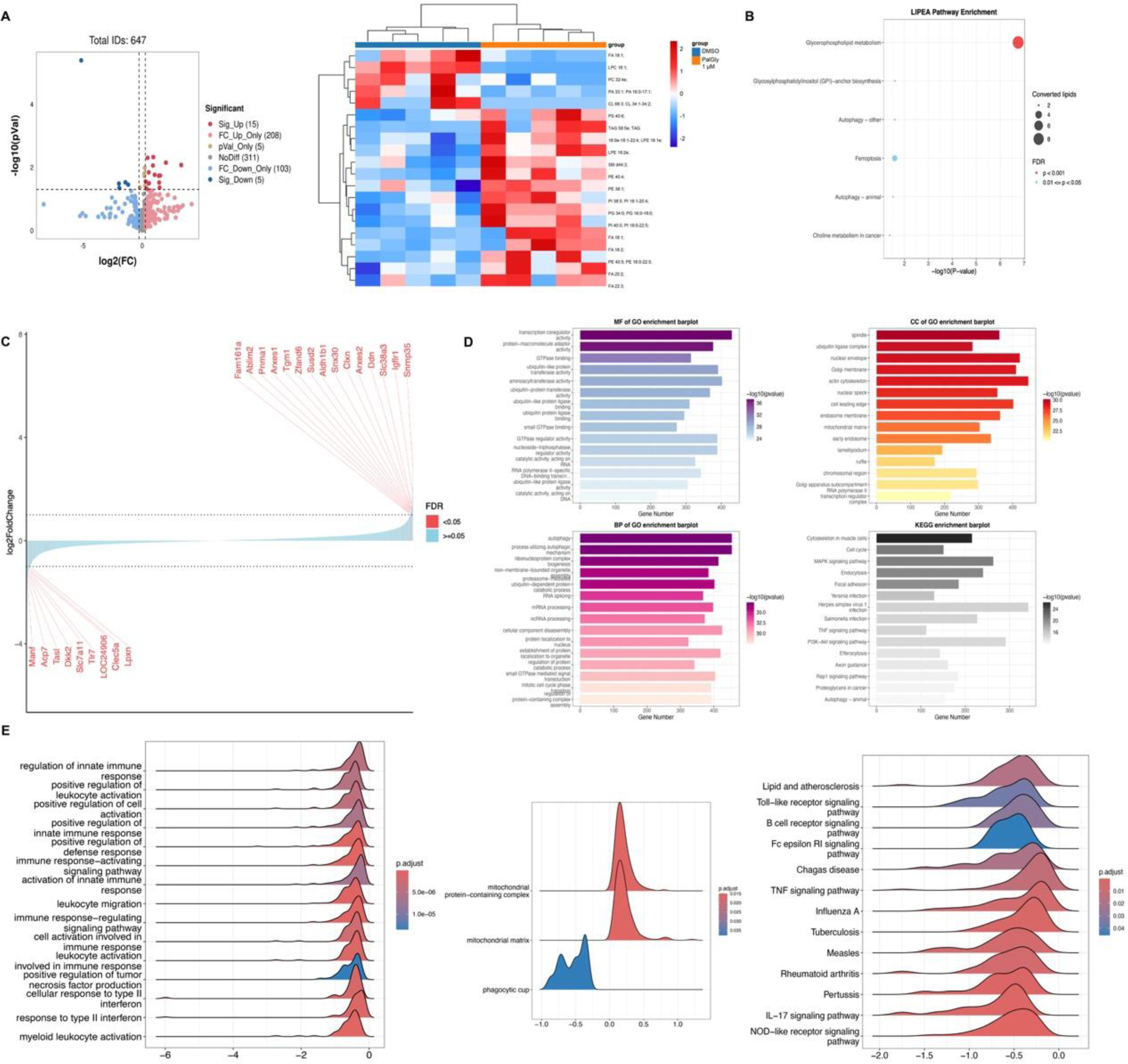
PalGly alters the transcriptomic and lipidomic profile of neonatal rat cardiomyocytes (A) Volcano plot and heatmap of differential lipids. (B) LIPEA lipids enrichment. (C) Significant differential expressed genes. (D) GO and KEGG enrichment. (E) GSEA analysis, GO categories are on the left and middle, and KEGG categories are on the right.

## 4.0 Discussion

Through conducting MR and colocalization analysis involving over 4000 proteins and 1000 metabolites, we have identified several previously unreported potential BrS targets as well as discovered the similarities and differences of these targets’ effects on BrS and other cardiometabolic disorders. Our MR analysis and colocalization analysis using eQTLs showed minimal overlap compared to the results by pQTLs. Given that proteins are regarded as primary function executors, we focused on investigating the proteome/metabolome-wide results. Combining the MR and colocalization results, we found that genetically predicted CXCL6 increased the risks of BrS and AF, and CR1 and DCC reduced the risks of BrS, leading to lower risks of cardioembolic stroke and HF respectively. PalGly and IGFBP3, on the other hand, increased the risk of BrS while reducing the risks of cardiometabolic diseases. Interestingly, molecular docking analysis revealed that PalGly not only has the potential to influence BrS risk itself but can also interact with BrS-related proteins (DCC, CR1, CTSB, NAAA, DEFB1, EPHA1, IGF1/IGFBP3/ALS, and LTA). Although mediation analysis indicated that only IGFBP3 exhibited a significant mediating effect. Furthermore, we identified for the first time that PalGly can enhance myocardial calcium sparks and shorten action potential duration (APD) and QT interval, with these electrophysiological effects likely mediated by the activation of TRPC5.

Although traditionally considered as a pure electrical disorder without structural alteration, there is accumulating evidence suggesting that myocarditis and inflammation infiltration may contribute to BrS pathogenesis ^38^, and C-reactive protein (CRP) was found to be an independent marker for symptomatic BrS^39^. Myocardial inflammation was detected in two patients with BrS and arrhythmic storm by fluorodeoxyglucose–positron-emission tomography (FDG-PET)^40^. One patient remained free of ICD shocks for 1 year after immunosuppression therapy. However, these studies were either cases or cross-sectional investigations, lacking robust and causal evidence. The CXCL6, CR1, and DCC screened by our analyses are proteins associated with immune response. CXCL6 primarily exerts pro-inflammatory effects by recruiting neutrophils^41^ and acts as a pro-angiogenic and pro-fibrotic paracrine factor secreted by endothelial cells and cardiac progenitor-like cells^41,42^. Elevated levels of CXCL6 are associated with higher idiopathic pulmonary fibrosis mortality^43^. CR1, on the other hand, exerts anti-inflammatory effects by facilitating the clearance of immune complexes and inhibits both C3 and C5 convertase activity^44^. Interestingly, administration of a recombinant form of soluble CR1 (TP10/CDX-1135) led to a decreased incidence of cardiovascular events in specifically male individuals undergoing cardiac surgery on cardiopulmonary bypass^45^. Gender difference is a well-recognized pattern of BrS, female patients with BrS are much rarer^46^, and the cohort included in this study is predominantly comprised of male participants (74%), suggesting inflammation may contribute to the gender difference of BrS. DCC is a netrin-1 receptor that is also involved in inflammation and angiogenesis and exerts its cardioprotective effect via Nitric oxide (NO) sustaining^47^. In-vitro evidence suggests that the cardioprotective effects exerted by netrin-1 in ischemia-reperfusion injury are by DCC-dependent endothelial-derived and cardiomyocyte-derived NO. Taken together, the immune/inflammation response is likely involved in the pathogenesis of BrS, and pharmacotherapies targeting these targets may hold promise as potential interventions.

Lipidomic and transcriptomic analyses indicated that PalGly negatively regulates the innate immune response, a finding that parallels the effects of TRPC5. PalGly is one of the fatty acid amides that serves as an endogenous signalling molecule^48^. It presents around 250-fold greater amounts in the skin (1612 pmol/g) compared to the amounts in the heart (6 pmol/g)^35^. Under the concentration of 27 μM, it potently inhibits heat-evoked firing of nociceptive neurons in rat dorsal horn and induced transient calcium influx in adult rat dorsal root ganglion (DRG) cells and a DRG-like cell line (F-11)^35^. Our results demonstrate that PalGly significantly increased the calcium spark amplitude in adult rabbit ventricular cardiomyocytes under field stimulation. Additionally, it augmented the width, duration, and frequency of calcium sparks while decreasing the time to peak. PalGly also significantly shortened APD both in vivo and ex vivo and shortened QT interval. We identified PalGly as an endogenous TRPC5 activator through patch-clamp experiments, the agonizing effect can be abolished by the TRPC5 inhibitor ML204. TRPCs contribute to Ca^2+^ signalling and changes in cell excitability. TRPC5 channels, like PalGly, are predominantly expressed in the brain^49^ and exist in a homomeric form or as a heteromer combined with TRPC1/TRPC4/TRPC5^50^. TRPC5 is a member of the TRP channel family, reportedly involved in cardiovascular pathophysiology^51^. Rosiglitazone, as a TRPC5 agonist, attenuates atrial structural remodelling and atrial fibrillation^52^, which is consistent with our findings that PalGly reduces the risk of AF. However, the underlying mechanism by which PalGly significantly shortens action potential duration (APD), and QT interval and ultimately increases the risk of BrS remains unclear. TRPC5 homomers and heteromers display distinct current-voltage (I-V) relationships. TRPC5 homomer or TRPC5/TRPC4 heteromer shows a doubly rectifying I-V curve characterized by substantial inward currents. In contrast, the TRPC5/TRPC1 heteromer exhibits predominantly outward rectification, manifesting relatively minor inward currents at negative potentials^53^. This distinct rectification behaviour may contribute to the changes observed in action potentials. Additionally, the intracellular calcium increase induced by TRPC5 activation can enhance the activity of calcium-dependent potassium channels (i.e. small conductance calcium-activated potassium channels, SK channels), promoting faster repolarization. PalGly may even directly activate relevant repolarization channels, contributing to these effects.

In addition, It was reported that IGFBP3 is significantly associated with higher sudden cardiac arrest (SCA) and syncope in the SCN5A mutation− BrS patients^54^ and higher serum calcium^55^. Our results showed that genetically predicted circulating IGFBP3 is associated with a higher risk of BrS, and the effect is partly mediated by the PalGly.

As BrS is relatively rare, we were unable to replicate our current findings through other cohorts. The cohorts available to us consisted of individuals of European ancestry, which may limit the generalizability of our results, particularly considering the higher prevalence of BrS in Southeast Asia^56^. We also were unable to perform sex-stratified and mutation-stratified analyses due to a lack of available summary-level statistics.

Collectively, our study underscores the involvement of PalGly, TRPC5, and inflammation-related proteins in the pathophysiology of BrS.

## Acknowledgements

L.W. is supported by the National Natural Science Foundation of China (No. 82370312 and 81930105). This work was supported by High-performance Computing Platform of Peking University. The funders had no role in the study design, data collection and analysis, decision to publish, or preparation of the manuscript. The authors thank members of all contributing studies for sharing their summary-level GWAS data.

## Author contribution

Conceptualization, H.X.; methodology, H.X., B.L.; software, formal analysis, and investigation, H.X.; electrophysiology experiments, H.X., B.L., and Y.C.; validation, H.X., B.L.; writing – original draft, H.X.; writing – review & editing, B.L., L.Y., A.Z., and L.W.; visualization, H.X.,

B.L.; funding acquisition, L.W.; supervision, L.W.

## Data availability

eQTLs: www.eqtlgen.org; deCode pQTLs: www.decode.com/summarydata/; UKB-PPP pQTLs: ukb-ppp.gwas.eu; MetabolitesQTLs: www.ebi.ac.uk/gwas/studies/GCST90199637; Brugada syndrome: www.ebi.ac.uk/gwas/studies/GCST90086158; ECGwas: http://www.ecgenetics.org; QTc interval: https://personal.broadinstitute.org/ryank/Nauffal_2022_QT_GWAS_SAIGE.zip; RNA-seq generated in this manuscript: GEO database under the code GSE278421.

## Code availability

Code for MR can be accessed at github.com/HongxuanXu/Multiomics-brugada-mr.

## Abbreviations

BrS: Brugada syndrome
ICD: implantable cardioverter-defibrillator
MR: mendelian randomization
ECG: electrocardiogram
GWAS: genome-wide association studies
QTL: quantitative trait loci
PBMC: peripheral blood mononuclear cell
IVW: Inverse variance weighted
ApoA1: apolipoprotein A1
ApoB: apolipoprotein B
HDL-C: high-density lipoprotein cholesterol
LDL-C: low-density lipoprotein cholesterol
Lp(a): Lipoprotein A
SBP: systolic blood pressure
DBP: diastolic blood pressure
HbA1c: Hemoglobin A1C
BMI: body mass index
AF: atrial fibrillation
CHD: coronary heart disease
HF: heart failure
PalGly: N-palmitoylglycine
APD: action potential duration
FDG-PET: fluorodeoxyglucose–positron-emission tomography
NO: Nitric oxide
DRG: dorsal root ganglion
DAD: delayed afterdepolarizations
SCA: sudden cardiac arrest

---

**Figure. S1.**
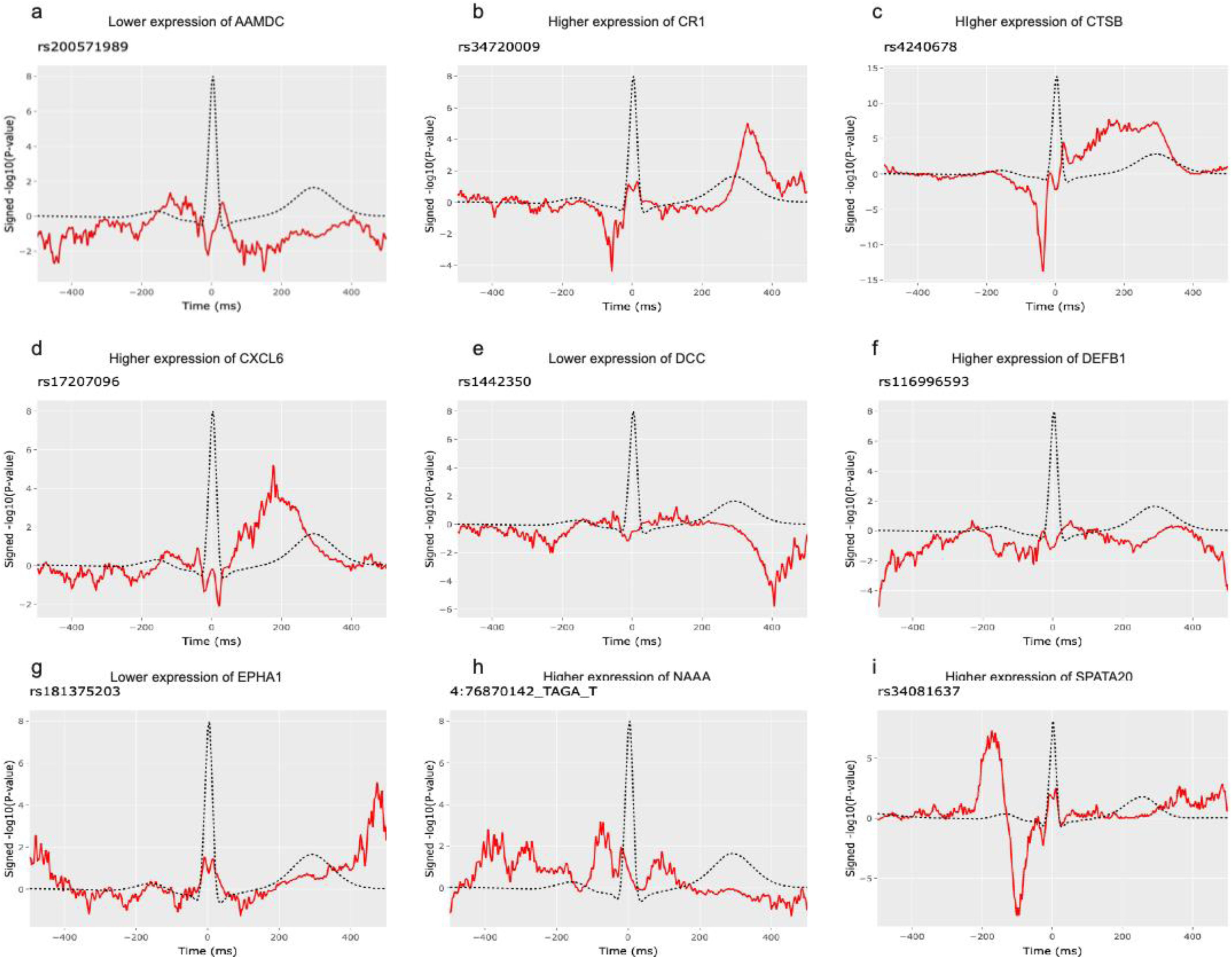
Circulating protein levels affect genetically proxied ECG morphology. Top cis-pQTLs were selected to represent the expression of the specific proteins

